# PREVALENCE OF ANEMIA, IRON DEFICIENCY ANEMIA, AND ASSOCIATED FACTORS AMONG DIABETICS IN THE WEST REGION of CAMEROON

**DOI:** 10.1101/2024.12.12.24318970

**Authors:** Leonard Tedong, Maï-Pamela Tumchou Mekieje, Melisa Mene Djeukamgang, Romaric Tuono De Manfouo, Marius Mbiandjeu Tchoumke, Pascal Blaise Well a Well a Koul, Josué Louokdom Simo, Pascal Dieudonne Chuisseu Djamen

**Affiliations:** Higher Institute of Health Sciences: Université des Montagnes, Bangangté, West-Cameroon; Biomedical Sciences, Faculty of Health Sciences: Université des Montagnes, Bangangté, West-Cameroon

**Keywords:** Anemia, iron deficiency anemia, diabetes, associated factors

## Abstract

**Aims and objective:** Anemias are a burden for management because they are associated with the worsening of the pathophysiology of diseases. They can be multifactorial, including micronutrient deficiencies including iron deficiencies. This study aimed to biologically characterize anemias and iron deficiency anemias during diabetes and to identify the associated factors.

**Methods:** Over 4 months, we conducted an analytical cross-sectional study from August to November 2024. 210 diabetic patients, attending the diabetology unit of the regional hospital in West Cameroon were studied, their age range between <20 and > 70 years. Blood samples were taken for biological tests including blood count, glycemia, glycated hemoglobin, ferritin, and serum following standard assay protocols. The results obtained were analyzed using the R statistical tool version 4.1.1.

**Results:** The sex ratio was 0.89 and the mean age was 48.32±18.19 years. 48.6% of patients were overweight. The frequency of anemia was observed in 72 females (53%) and 63 males (46.7%) with a total frequency of 64.3%. 11.4% were found to have iron deficiency anemias. The blood count revealed hematological abnormalities characterized by leukopenia (41.4%), granulocytopenia (35.7%), and thrombocytopenia (2.86%) with macroplateletosis (40.0%). Anemias were characterized by microcytosis (11.1% in anemic patients versus none in non-anemic patients (*p*<0.001), hypochromia (88.9% in anemic patients versus 72% in non-anemic patients (*p*<0.001)) and greater regeneration in anemic patients versus non-anemic patients (p<0.001). Macroplateletosis and high Hb1AC were identified as predictive factors for iron deficiency anemia in diabetes with (Or=2.83, 95% CI= [1.17;6.80]; *p*=0.021) and Or=2.38, 95% CI= [0.14;3.09], *p*=0.045, respectively.

**Conclusion:** This study reports a significant frequency of anemia and iron deficiency anemia in diabetic patients; the latter is associated with worsening clinical manifestations in patients. This study raises the need for regular exploration of iron status in diabetics to prevent complications related to iron deficiency.

## 1. INTRODUCTION

Anemia is defined as a condition in which the number and size of red blood cells or the concentration of hemoglobin falls below normal values [1]; resulting in a lack of oxygenation of the tissues of the human body [2]. Current recommendations for the threshold of hemoglobin in the blood range from 13 to 14.2 g/dL in men and 11.6 to 12.3 g/dL in women [3]. The World Health Organization estimates that approximately one quarter of the world’s population suffers from anemia [3]. Anemia is estimated to affect half a billion women aged 15 to 49 years and 269 million children aged 6 to 59 months [3]. The African region and the Asian region are the most affected with 106 million women and 103 million children affected by anemia in Africa and 244 million women and 83 million children affected in South East Asia [3]. Anemia can come from various causes, namely iron deficiency, vitamin B12 deficiency, folate deficiency [4]. According to the WHO, Iron Deficiency Anemia (IDA) is the most widespread form in the world affecting approximately 2 billion people or 30% of the world population [3]. Indeed, Iron Deficiency Anemia (IDA) is a global public health problem that affects both developing and developed countries, with major consequences on human health as well as on social and economic development [5]. Furthermore, given the magnitude of the problem, several consequences can result from it, for example, annoying symptoms and reduced quality of life; and affecting all aspects of physical and emotional well-being [6,7]. These iron deficiency anemias are the states that can be encountered during certain diseases including genetic disorders, bacterial and parasitic infections, and metabolic diseases including diabetes [5].

Indeed, diabetes is a metabolic disease characterized by a chronic elevation of the blood glucose level (hyperglycemia) in relation to peripheral resistance to insulin (type II diabetes) or a deficiency in its production (type I diabetes) [8]. It is a disease in perpetual growth in the world (480 million cases in 2014 compared to 536.6 million in 2021) [9]. In Cameroon, more than three million of the population is diabetic [10]. Anemia in diabetic patients seems to have a remarkable adverse effect on the quality of life and is associated with the progression of the disease and the development of comorbidities [11]. Iron deficiency anemia appears to be more common in diabetic patients than in non-diabetics especially those with nephropathy [11]. Iron deficiency can impair glucose homeostasis in humans negatively affect glycemic control, and predispose to more complications in diabetic patients [12]. Studies have reported that iron deficiency anemia contributes to the aggravation of diabetes-related diseases including nephropathy, retinopathy, manifestations of ischemic heart disease, and left ventricular hypertrophy and in extreme cases could lead to a reduction in the half-life of patients [12].

Diabetes is a public health problem because of the intrinsic complications that patients are prone to [11–13]. Associated with this, nutritional deficiencies including iron deficiency are further factors contributing to the aggravation of the pathological state of patients. Moreover, anemia is a real burden whose poor management can be the cause of multiple complications. However, iron deficiency anemia in general and iron deficiency anemia in particular are little or poorly studied in the world in general and in countries with limited resources such as Cameroon. However, the latter could have harmful consequences among which they could contribute to affecting the glycemic control of patients [12,14–16]. The clinical relevance of the effect of iron deficiency on glucose metabolism is not yet clear. The links between glucose, anemia, and HbA1c are complex and not yet completely elucidated. This study therefore aims to determine and diagnose iron deficiency anemia in Cameroonian diabetic patients, to biologically characterize these anemias to provide some clarification on the subject in particular in diabetic patients as part of the control and prevention of the consequences associated with poor anemic control in diabetics.

## 2. METHODS

### 2.1. Ethical considerations

This study was conducted after obtaining study authorizations and ethical clearance issued by the Institutional Ethics and Research Committee of the University of the Mountains (N°. 2024/211/UdM/PR/CEAQ) on 31 July 2024. The authorization for collection and analysis (N°. 663/L/MINSANTE/SG/DRSPO/HRB/D) was obtained by the Regional Hospital of Bafoussam on 07 August 2024. Participants in this study were informed about the benefits and risks of the research, the progress, and the objective of the study through a questionnaire. Then, they provided written consent before they were recruited into the study. To ensure the confidentiality of the information obtained from the participants, an anonymity number was assigned to each participant. This study was conducted in strict compliance with scientific and medical research as set out in the Declaration of Helsinki [17].

### 2.2. Study design, study location and study population recruitment strategy

The study conducted was an analytical cross-sectional study over 04 months and data collection took place between August 07, 2024 and November 30, 2024. The Bafoussam Regional Hospital (HRB) served as a framework for the recruitment of participants in this study. Indeed, according to the health pyramid of Cameroon, the Bafoussam Regional Hospital is a third category and second reference public hospital created in 1953. It treats common diseases suffered by the population, including metabolic diseases including diabetes. It is located in the West Cameroon region, Mifi department. The data collection from the participants was carried out in its diabetology unit. The diabetology center of the Bafoussam Regional Hospital is a center dedicated to the care and monitoring of diabetic patients. The biological analyses of the blood samples were carried out at the Medical Analysis Laboratory of the said hospital for the hematology part and at the Biochemistry laboratory of the University of the Mountains for the second part.

An estimate of the size of the study population was made from a study conducted by Kabamba et *al*. (2015) on anemia in diabetic patients in the Congolese population reporting 57% anemia in diabetics [18].

For each participant, clinical, biological, sociodemographic and anthropometric data were obtained from a pre-established form including the date of diagnosis of the disease, the type of diabetes, family history of diabetes, type of treatment, lifestyle habits, comorbidities. Patients treated in the said Center who had given their informed consent and were able to provide the necessary and useful information for the study were included. Those unable to provide it and pregnant or breastfeeding women were not included. A total of 210 diabetic patients were included in the study.

#### Procedure for collecting data and biological analyses of samples

After filling out the questionnaire and obtaining anthropometric data, a volume of blood was collected in 02 tubes for hematological analyses and others biochemical parameters.

Concerning the carrying out of hematological analyses, it was done from the realization of the hemogram using a hematology machine (MINDRAY BC 2800) using the principle of cytometry for counting blood cells. Then, the results obtained were analyzed and interpreted according to the reference values defined by the WHO. Subsequently, the confirmation and classification of cases of anemia, and a better assessment of the quality of the cells was made thanks to the realization of the reticulocyte count; this from a blood smear stained according to the Rovanowsky principle [19]. The slides were read using an optical microscope of the model: Olympus CX23.

Concerning the performance of biochemical analyses, the assessment of the patient’s iron status was made from the determination of ferritinemia by immunoenzymatic method using the ELISA Indirect principle according to the protocol of the reagent ’Fortress Diagnostics’ of the batch number BX0891. Then, serum iron was determined by spectrophotometric method using FerroZine reagent.

Glycemic management was assessed from the dosage of glycated hemoglobin (HbA1c) by TINIA type immunoturbidimetric method (Turbidimetric inhibition immunoassay) carried out on hemolyzed whole blood. Glycemic status was assessed from the measurement of blood sugar in a punctual manner by spectrophotometric method according to the protocol defined by Biolabo.

### 2.3. Data management and statistical analysis

The data shown below were entered into a Microsoft Excel 2016 spreadsheet and analyzed using the R statistical tool version 4.1.1. The variables analyzed were the quantitative variables: serum iron, ferritin, HbA1c, MCV, MCHC, MCHC, platelet, MPV, hemoglobin level, red blood cells, white blood cells, platelets, reticulocytosis, MCV, MCHC, blood glucose; qualitative variables: age groups, sex, BMI, clinicobiological data. Quantitative variables were categorized into ‘low, normal, high’ according to the reference values defined by the manufacturer or WHO where applicable. For this study, anemia was defined as hemoglobin values <12g/dL in women and 13g/dL in men. Iron deficiency was defined as ferritin levels < 30ng/mL in men and < 15ng/mL in women[20]. Poor glycemic control was defined as glycated hemoglobin values >7%, poor blood sugar levels defined as values below 1.20g/L [21]. Iron deficiency anemia was defined as patients with both anemia and iron deficiency [5].

A normality test was performed to ensure the normal distribution of values. The results were presented as mean and standard deviation for quantitative variables and frequencies for qualitative variables. The student test was used to compare quantitative variables while Pearson’s chi2 test allowed the comparison of proportions. A 95% confidence interval whenever necessary. Logistic regression analyses according to the univariate model allowed us to identify the factors associated with anemia and deficiency anemia. The significance threshold will be set for values of *p* < 0.05.

## 3. RESULTATS

210 diabetics were included in this study. Of these, 135 patients (64.3%) presented anemia. The mean age of the study population was 48.2±20.3 years.

### 3.1. Distribution of the study population according to socio-demographic, anthropometric and clinical history characteristics

#### 3.1.1. Distribution of the study population according to socio-demographic and anthropometric data

The table below describes the socio-demographic characteristics and anthropometric data of the study population according to anemia:

Table 1 above shows that gender does not differ significantly according to anemia in diabetic patients in this study (p = 0.97) with a female predominance (52.9%). The most represented age group was >70 years with 39 (18.6%) patients. Although not differentiating according to the presence of anemia (p>0.05), 48.6% of patients were overweight and 17.1% obese.

**Table 1:**
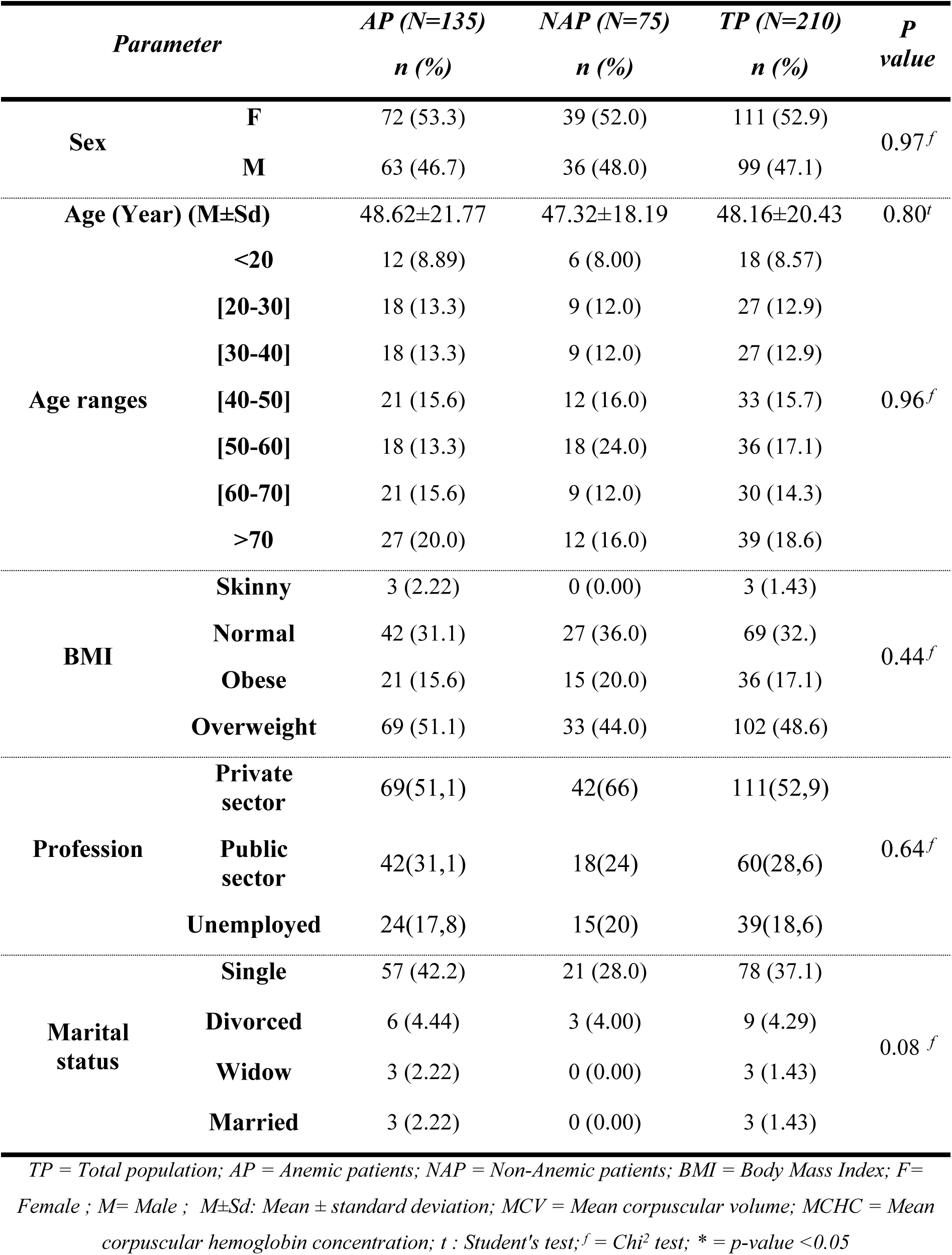
Sociodemographic and anthropometric characteristics of the study population according to anemia.

#### 3.1.2. Distribution of the study population according to clinical and biological history

Table 2 below presents the clinical and biological history of the diabetic patients in this study:

**Table 2:**
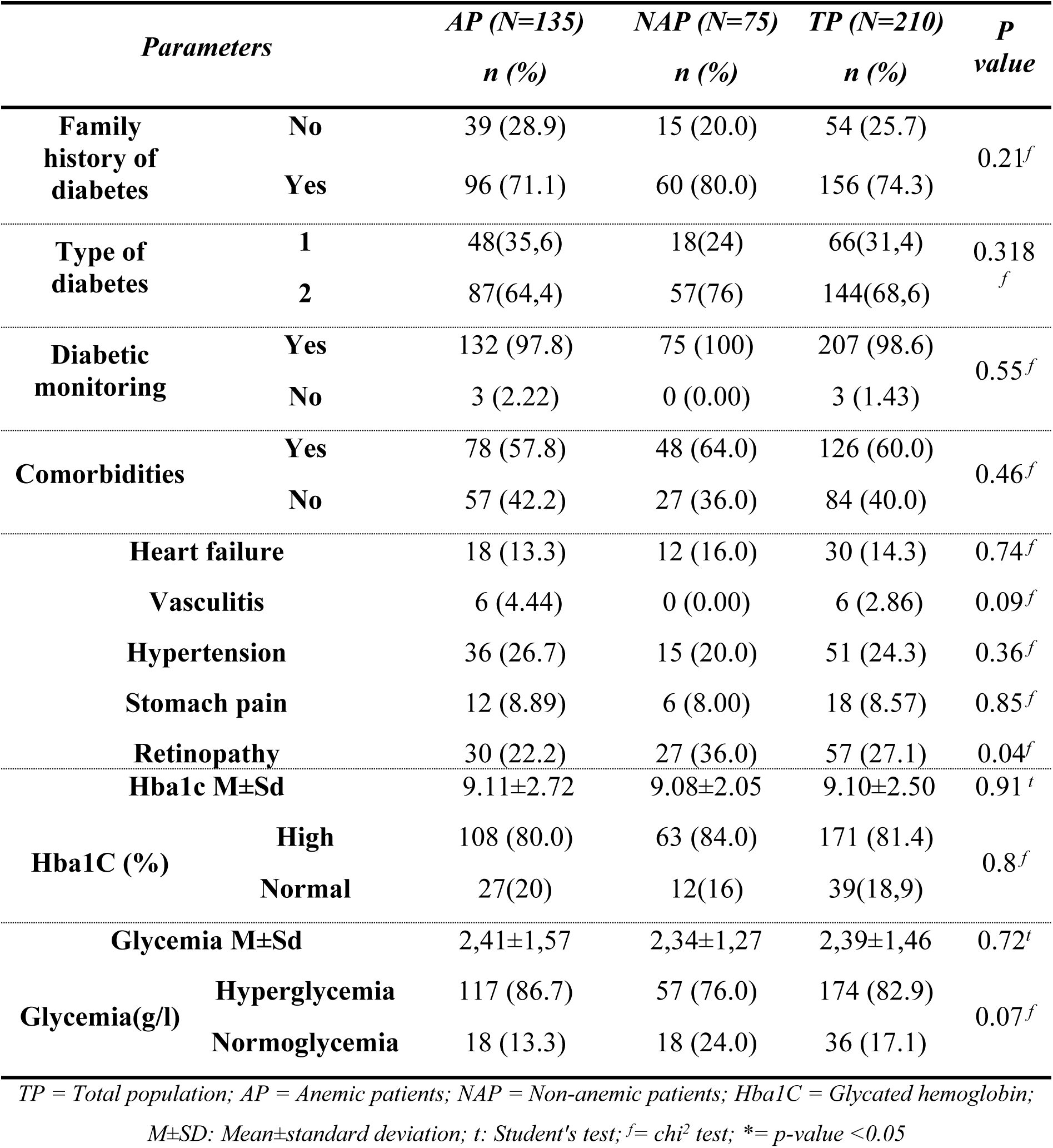
Distribution of the study population according to clinical and biological history.

From Table 2 above, type 2 diabetes was the most represented (68.6%) in the population; and the type of diabetes did not significantly differentiate according to the presence or absence of anemia (p>0.05). The study population had a family history of diabetes (74.3%). Almost the entire population (98.6%) studied was undergoing diabetic follow-up. The patients studied had hyperglycemia (82.9%) and elevated Hba1c (81.4%) although these parameters did not differentiate according to the presence of anemia. The study population had comorbidities characterized by the presence of vasculitis (2.86%), high blood pressure (24.3%), retinopathy (27.1%), gastric pain (8.57%), heart failure (14.3%).

### 3.2. Population distribution according to biological data

#### 3.2.1. Blood count profile in the population among diabetics

##### 3.2.1.1. Red lineage profile, anemia and biological characterization

Table 3 below describes the red lineage profile and biological characterization of anemias in diabetic patients:

**Table 3:** Red lineage profile and biological characterization of anemias in the population.

| *Parameters* |  | *AP (N=135)* | *NAP (N=75)* | *TP (N=210)* | *P value* |
| --- | --- | --- | --- | --- | --- |
|  |  | *n (%)* | *n (%)* | *n (%)* |  |
| **RBC (T/L) M±Sd** |  | 4.38±0.74 | 4.81±0.54 | 4.53±0.70 | <0.001<sup>t</sup> |
| **RBC** | **Low** | 30 (22.2) | 6 (8.00) | 36 (17.1) | 0.003<sup>ft</sup> |
|  | **High** | 6 (4.44) | 0 (0.00) | 6 (2.86) |  |
|  | **Normal** | 99 (73.3) | 69 (92.0) | 168 (80.0) |  |
| **HB(g/dL) M±Sd** |  | 10.2±0.93 | 12.5±0.99 | 11.0±1.46 | <0.001<sup>f*</sup> |
| **HB** | **Low** | 135 (100) | 0 (0.00) | 135 (64.3) | <0.001<sup>f*</sup> |
|  | **Normal** | 0 (0.00) | 75 (100) | 75 (35.7) |  |
| **MCV M±Sd** |  | 84.8±13.6 | 90.0±4.60 | 86.6±11.5 | <0.001<sup>f*</sup> |
| **VGM (fL)** | **Low** | 15 (11.1) | 0 (0.00) | 15 (7.14) | 0.001<sup>f*</sup> |
|  | **Normal** | 117 (86.7) | 75 (100) | 192 (91.4) |  |
|  | **High** | 3 (2.22) | 0 (0.00) | 3 (1.43) |  |
| **MCHC M±Sd** |  | 27.0±2.69 | 27.4±1.69 | 27.2±2.39 | 0,26<sup>t</sup> |
| **MCHC**<br>**(g/dL)** | **Low** | 132 (97.8) | 75 (100) | 207 (98.6) | 0.55<sup>f</sup> |
|  | **Normal** | 3 (2.22) | 0 (0.00) | 3 (1.43) |  |
| **MCH M±Sd** |  | 23.7±2.92 | 25.2±2.21 | 24.2±2.77 | <0.001<sup>t*</sup> |
| **MCH (pg)** | **Low** | 120 (88.9) | 54 (72.0) | 174 (82.9) | 0.003<sup>f*</sup> |
|  | **Normal** | 15 (11.1) | 21 (28.0) | 36 (17.1) |  |
| **Hématocrit (M±Sd)** |  | 38.1±5.84 | 45.3±5.15 | 40.6±6.57 | <0.001<sup>t*</sup> |
| **Hématocrit**<br>**(%)** | **Low** | 123 (91.1) | 42 (56.0%) | 165 (78.6) | <0.001<sup>t*</sup> |
|  | **Normal** | 9 (6.67) | 24 (32.0%) | 33 (15.7) |  |
|  | **High** | 3 (2.22) | 9 (12.0%) | 12 (5.71) |  |
| **Rt (G/L) M±Sd** |  | 117±41.9 | 79.4±36.5 | 104±43.9 | <0.001<sup>t*</sup> |
| **Rt** | **Low** | 0 (0.00) | 21 (28.0) | 21 (10.0) | <0.001<sup>f</sup> |
|  | **Normal** | 135 (100) | 54 (72.0) | 189 (90.0) |  |

*RBC = Red Blood Cells; HB = Hemoglobin; Rt = Reticulocytes; TP = Total Population; AP = Anemic Patients; NAP = Non-Anemic Patients; M±SD: Mean ± Standard Deviation; MCV = Mean Cell Volume; MCHC = Mean Corpuscular Hemoglobin Concentration; G/L=Gigal/Liter; T/L=Tera/Liter; MCH: Mean Corpuscular Hemoglobin Content; Rt = Reticulocytes; t: Student's test; f = Chi2 test; \* = p-value <0.05*

The results show a mean red blood cell count of 4.53 T/L ± 0.71 in the total population including 4.33 T/L ± 0.74 in Anemic Patients (AP) and 4.81 T/L ± 0.54 in Non-Anemic Patients (NP) (*p* = 0.01). Hemoglobin levels show a mean of 10.2 ± 0.93 g/dL in AP, and 12.5 ± 0.99 g/dL in NAP (*p* < 0.001). MCV showed a significant difference in anemic patients compared to non-anemic patients (p<0.001) characterized, with a mean of 84.8 ± 13.8 fL in AP and 90.0 ± 4.6 fL in NAP (*p*<0.001) including 11.1% microcytosis in AP versus none in NAP (*p*<0.001). Hypochromia was significantly higher (p<0.001) in AP (88.9%) compared to PNA (72.0) characterized by a mean MCH of 23.7±2.92 pg in AP and 25.2±2.21 pg in NAP (*p*<0.001). The mean reticulocyte rate in AP is 117±41.9 G/L and 79.4±36.5 G/L in NAP (*p*<0.001) characterizing an ageneration 8 in AP (*p*<0.001).

##### 3.2.1.2. White and platelet lineage profile

Table 4 below describes the white blood cell and platelet profile in diabetic patients:

**Table 4:**
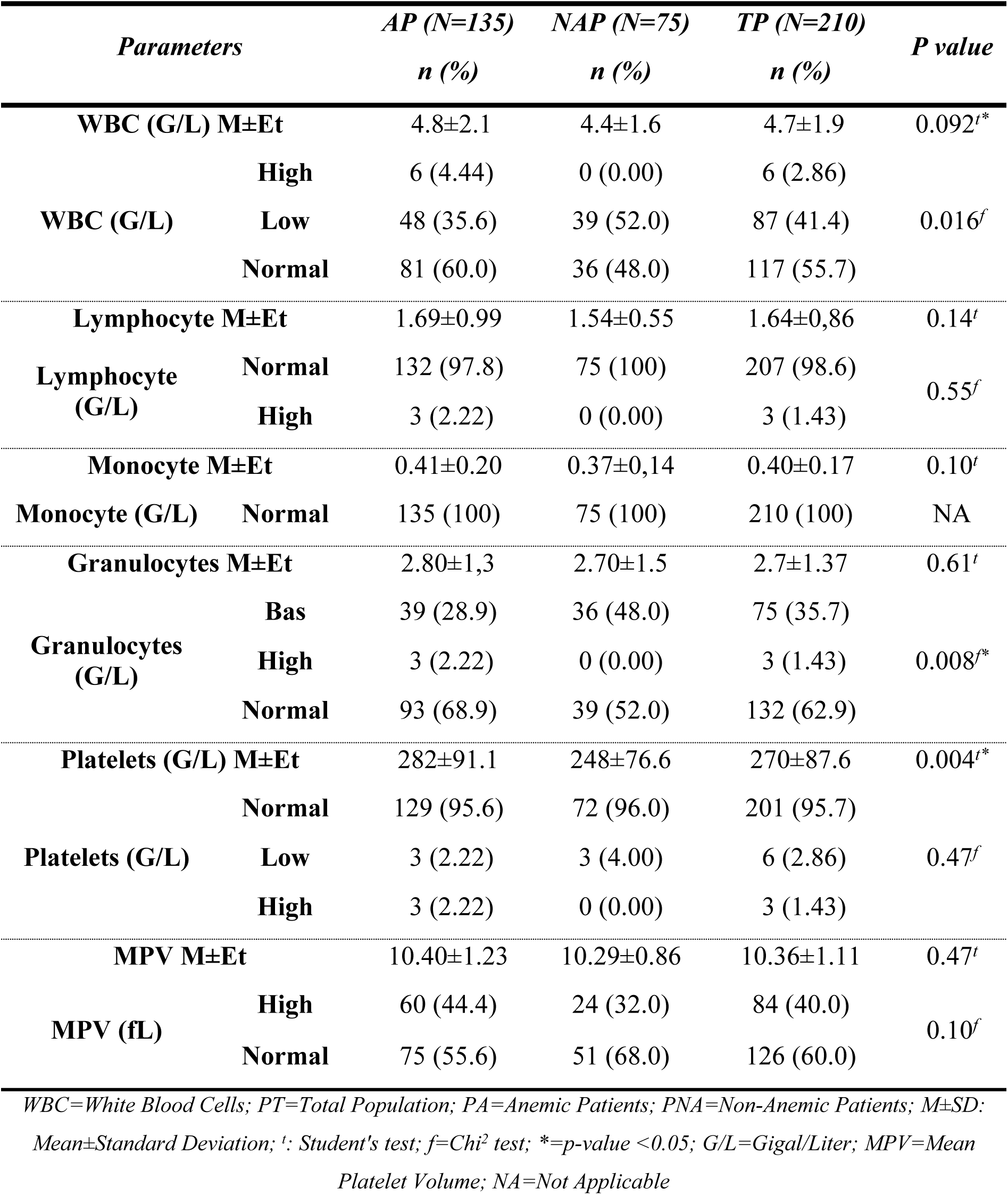
White blood cell and platelet profile in the population.

From the table above it is clear that 41.4% of patients had leukopenia. 35.7% had granulocytopenia and 40% had macroplatelets.

#### 3.2.2. Frequency of iron deficiency and iron deficiency anemia, iron parameters in the population and associated factors

##### 3.2.2.1. Frequency of iron deficiency and iron deficiency anemia

At the end of the study, 33 (15.7%) patients presented iron deficiency but it did not differ according to the presence of anemia. Of these, 24 (11.4%) presented iron deficiency anemia. Table 5 below shows the distribution of the population according to iron parameters and iron deficiency in the population according to anemia.

**Table 5:**
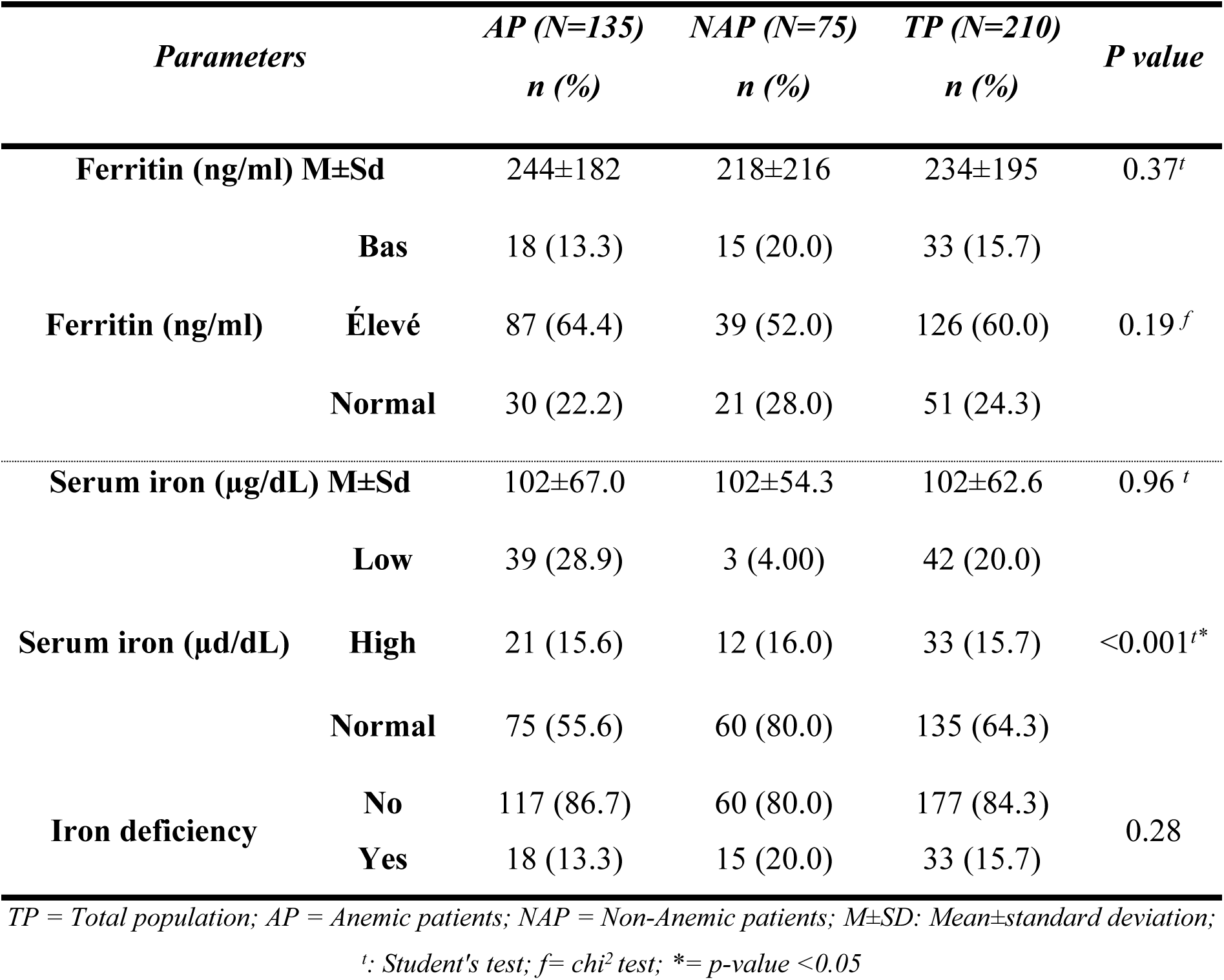
Iron parameters and iron deficiency in the population.

From Table 5 above it is clear that 33 (15.7) patients presented hypoferritinemia. Furthermore, anemic patients presented a significant decrease in serum iron compared to NAP (p <0.001).

##### 3.2.2.3. Factors associated with anemia

###### 3.2.2.3.1. Factors associated with anemia during this study

The following Table 6 describes the factors associated with anemia in the population

**Table 6:** Factors associated with anemia during this study: univariate analysis.

| *Parameters* | *Anemic (N=135)*<br>*n (%)* | *Non anemic*<br>*(N=75)*<br>*n (%)* | *OR [IC 95%]* | *P-value* |
| --- | --- | --- | --- | --- |
| **Sex:** |  |  |  |  |
| **F** | 72 (53.3) | 39 (52.0) | Ref. | Ref. |
| **M** | 63 (46.7) | 36 (48.0) | 0.95 [0.54;1.67] | 0.854 |
| **Hb1AC** |  |  |  |  |
| **High** | 108 (80.0) | 63 (84.0) | 2.53 [0.59;3.15] | 0.0048 |
| **Normal** | 27 (20) | 12 (16.0) | Ref. | Ref. |
| **Granulocytes:** |  |  |  |  |
| **Low** | 39 (28.9) | 36 (48.0) | Ref. | Ref. |
| **High** | 3 (2.22) | 0 (0.00) | 2.20 [1.22;3.96] | 0.009 |
| **Normal** | 93 (68.9) | 39 (52.0) | . [.;.] | 0.151 |
| **Ferritin:** |  |  |  |  |
| **Low** | 18 (13.3) | 15 (20.0) | Ref. | Ref. |
| **High** | 87 (64.4) | 39 (52.0) | 1.86 [0.85;4.07] | 0.128 |
| **Normal** | 30 (22.2) | 21 (28.0) | 1.19 [0.49;2.88] | 0.705 |
| **Iron Deficiency:** |  |  |  |  |
| **No** | 117 (86.7) | 60 (80.0) | Ref. | Ref. |
| **Yes** | 18 (13.3) | 15 (20.0) | 0.62 [0.29;1.31] | 0.214 |
| **Iron Deficiency Anemia:** |  |  |  |  |
| **No** | 126 (93.3) | 60 (80.0) | Ref. | Ref. |
| **Yes** | 9 (6.67) | 15 (20.0) | 0.29 [0.12;0.69] | 0.005 |

*F=female; M=Male; Or= Odd ratio; 95% CI = 95% Confidence Interval; Hb1AC= Glycated hemoglobin; significance test $p<0.05$*

It is clear from the table above that granulocytosis and hyperferritinemia were identified as predictive factors for anemia with respectively Or=2.20 IC95%=[1.22;3.96]; *p*=0.009 and Or=1.86 IC 95%= [0.85;4.07]; *p*= 0.128.

###### 3.2.2.3.2. Factors associated with Iron Deficiency Anemia

The following Table 7 describes the factors associated with iron deficiency anemia in the population

**Table 7:** factors associated with iron deficiency anemia in the population, univariate analyses.

| *Parameters* | *Absence of IDA*<br>*(N=186)*<br>*n (%)* | *Presence of IDA*<br>*(N=24)*<br>*n (%)* | *OR [IC 95%]* | *p-value* |
| --- | --- | --- | --- | --- |
| **Sex:** |  |  |  |  |
| **Female** | 102 (54.8) | 9 (37.5) | Ref. | Ref. |
| **Male** | 84 (45.2) | 15 (62.5) | 0.49 [0.21;1.19] | 0.117 |
| **Glycemia:** |  |  |  |  |
| **Hyperglycemia** | 153 (82.3) | 21 (87.5) | 1.51 [0.43;5.36] | 0.558 |
| **Normoglycemia** | 33 (17.7) | 3 (12.5) | Ref. | Ref. |
| **Hba1C, Mean±sd** | 9.19±2.56 | 8.37±1.86 | 1.14 [1.36;0.96] | 0.132 |
| **Hba1C** |  |  |  |  |
| **High** | 156 (83.9) | 15 (62.5) | 2.38 [0.14;3.09] | 0.045 |
| **Normal** | 30 (16.1) | 9 (27.5) | Ref. | Ref. |
| **Hemoglobin, Mean±sd** | 11.0±1.47 | 11.0±1.43 | 1.00 [1.34;0.74] | 0.984 |
| **Hemoglobin:** |  |  |  |  |
| **Anemia** | 126 (67.7) | 9 (37.5) | Ref. | Ref. |
| **Normal** | 60 (32.3) | 15 (62.5) | 0.29 [0.12;0.69] | 0.005 |
| **MCH, Mean±sd** | 24.4±2.76 | 23.2±2.73 | 1.14 [1.31;0.99] | 0.068 |
| **MCH:** |  |  |  |  |
| **Hypochromia** | 153 (82.3) | 21 (87.5) | Ref. | Ref. |
| **Normochromia** | 33 (17.7) | 3 (12.5) | 1.51 [0.43;5.36] | 0.558 |
| **Granulocytes:** |  |  |  |  |
| **Low** | 63 (33.9) | 12 (50.0) | Ref. | Ref. |
| **High** | 3 (1.61) | 0 (0.00) | . [.;.] | 0.602 |
| **Normal** | 120 (64.5) | 12 (50.0) | 1.90 [0.81;4.48] | 0.149 |
| **MPV:** |  |  |  |  |
| **Macroplateletosis** | 69 (37.1%) | 15 (62.5) | 2.83 [1.17;6.80] | 0.021 |
| **Normoplateletosis** | 117 (62.9%) | 9 (37.5) | Ref. | Ref. |
| **Ferritin, Mean±sd** | 263±189 | 13.5±7.60 | 1.15 [1.23;1.08] | <0.001 |

*F=female; M=Male; Or= Odd ratio; 95% CI = 95% Confidence Interval; Hb1AC= Glycated hemoglobin; IDA =Iron Deficiency Anemia ; significance test $p<0.05$*

From this table, gender is not a factor associated with iron deficiency anemia in diabetes. In addition, macroplateletosis and high Hb1AC were identified as predictive factors of iron deficiency anemia in diabetes with respectively (Or=2.83, 95% CI=[1.17;6.80]; *p*= 0.021) and Or=2.38, 95% CI=[0.14;3.09], *p*=0.045.

## 4. DISCUSSION

The aim of this study was to determine the frequency of anemia and iron deficiency anemia; and to biologically characterize the latter during diabetes. Indeed, anemia is a situation commonly encountered in diabetics. It is most often the consequence of complications of diabetes, in particular kidney disease, but it can be the consequence of treatments, in particular, renin-angiotensin system inhibitors, antiplatelet agents, metformin[22]. They have several mechanisms, including inflammatory and non-inflammatory, and/or iron deficiency [23]. Iron deficiency anemia is the most common micronutrient deficiency and is the third leading cause of anemia worldwide [5,15]. Thus, the frequency of anemia reported during this study was 64.3%; that of iron deficiency anemia was 11.4%. Kabamba et *al*. (2015) in Congo reported 57% of anemia[18] ; Ebrahim et *al*. (2022 ) in Ethiopia reported 25.8% anemia [24] ; Kim et *al*. (2021) reported 11.29% anemia in Korea[15], Aparecida et *al.* (2021) [25] in Brazil reported 34.2%), Tujuba et *al.* in Ethiopia reported 30.2% [26], and Sharif et al. rapportaient 2014 rapportaient 63% au Pakistan [27]. Moreover, Christy et *al.* (2014) reported 32.9% iron deficiency anemia in diabetics [28]. Indeed, diabetes can contribute to anemia and iron deficiency by reducing iron absorption, causing gastrointestinal bleeding, and causing diabetic complications [11,29]. These different studies report different frequencies of anemia during diabetes, which could be explained by differences that could have existed in the different contexts in which each of these studies were carried out; however, they still raise a significant frequency of anemia and iron deficiencies during diabetes.

Type 2 diabetes was the majority in our population 68.6%. This could be explained by the fact that type two diabetes increases with age and constitutes middle-aged diabetes and is the most prevalent diabetes in the world according to WHO [30]. The mean age of the study population was 48.16±20.43 years with the age group >70 years being the largest (18.6%). Indeed, diabetes and in particular type 2 diabetes is progressive in onset [13]. Several authors also report an advanced age during diabetes [31,32]. A majority of the population was overweight 48.6%; 17.1% were obese. This could be explained by the fact that obesity is a risk factor for diabetes [31]. The treatments that the study participants were following were mainly those of the class of oral antidiabetics in which are cited among others metformin, Diamicron, glinide, Amarel. These constitute the first line of medication used in antidiabetic treatment and constitute the oral antidiabetics recommended as first-line or in combination with any other antidiabetic treatment for the treatment of type 2 diabetes and the pleiotropic properties of metformin suggest that this antidiabetic acts simultaneously on multiple tissues by different mechanisms to control glycemia [27] ; This has also been reported by several authors [12,33– 35]. Such approaches play a vital role in preventing and delaying the onset and progression of diabetic complications including heart failure, ocular involvement, gastric pain, vasculitis nephropathy; which correspond to the comorbidities reported in the literature as commonly encountered in diabetics [5,26,36].

Furthermore, HbA1c reported during this study, although high in the general population, did not present a significant difference on the one hand according to the presence or absence of anemia, and on the other hand according to the presence or absence of anemia by iron deficiency. According to the pharmacological management of DM, HbA1c should be assessed regularly in patients with diabetes. The frequency of HbA1c is flexible and depends primarily on the response of patients to therapy and physician judgment. Indeed, there are several controversies in the literature regarding the influence of glycated hemoglobin on iron deficiency. Although, we have identified a high Hb1AC as a predictive factor of anemia by iron deficiency during diabetes with respectively (Or = 2.83, CI 95% = [1.17; 6.80]; *p* = 0.021); and that an increase in the level of HbA1C is associated with anemia; which was also noted by Christy et *al*. found a positive correlation between IDA (patients with Hb: = 9.4 ± 1.3 g/dL) and increased A1C levels, [28] and several authors also report that Increased Levels of Glycated Hemoglobin A1c and Iron Deficiency Anemia [16,37]. However, previously published studies have shown that the effect of iron deficiency and iron deficiency anemia on HbA1c levels occurs independently [38,39]. Attard et *al*. studied HbA1c levels and fasting blood glucose as diagnostic criteria to assess the effect of iron deficiency and iron deficiency anemia on diabetes. The results showed that iron deficiency and iron deficiency anemia caused changes in HbA1c levels, which were not consistent with the actual disease status [39]. Moreover, anemia is characterized by a shorter erythrocyte lifespan, reduced hemoglobin concentration and compensatory hyperplasia [40]. All these factors exert significant changes in HbA1c production. Iron deficiency has been reported to be independently associated with increased HbA1c, independent of plasma glucose levels and degree of anemia [14,41]. Studies have shown a significant reduction in HbA1c in non-diabetic patients with controlled plasma glucose levels after administration of iron supplements [34–36]. Furthermore, other studies have shown no correlation between markers of iron storage (ferritin) and increased HbA1c. [28].

The exploration of the blood count showed a general alteration of hematological parameters characterized by leukopenia (41.4%), granulocytopenia (35.7%), 40.0% of macroplateletosis, the latter of which was identified as a predictive factor of iron deficiency (Or = 2.83, 95% CI = [1.17; 6.80]; *p* = 0.021). Ebrahim et *al*. (2022) also reported a general alteration of hematological parameters during diabetes; as well as several other authors [24,42]. Indeed, it has been reported that prolonged hyperglycemia can cause an increase in the production of reactive oxygen species (ROS) and the formation of significant advanced glycation end products (AGEs); and these are directly associated with endothelial dysfunction and hematological changes. Moreover, acute activation of oxidative stress caused by excessive ROS release can lead to tissue damage and hematological alterations such as red blood cell dysfunction, platelet hyperactivation, and endothelial dysfunction [43]. In addition, insulin resistance is also associated with endothelial dysfunction, hyperactivation and increased plasma levels of inflammatory markers, leukocytosis and platelet hyperactivation that can trigger and accelerate vascular complications in patients with type 2 diabetes in particular [37,44]. On the other hand, leukopenia and granulocytopenia would reflect a decrease in immune defenses during diabetes resulting from the comorbidities to which patients are subject. [24]

Concerning anemias, they were mainly characterized by microcytoses with a mean MCV of 84.8 ± 13.8 fL in AP and 90.0 ± 4.6 fL in NAP (*p* < 0.001) and including 11.1% of microcytosis in AP versus none in PNA (p < 0.001); significantly higher hypochromias (*p* < 0.001) in AP (88.9%) compared to NAP (72.0) characterized by a mean TCMH of 23.7 ± 2.92 pg in AP and 25.2 ± 2.21 pg in NAP (*p* < 0.001); and mainly aregenerative anemias. On the other hand, iron deficiency in this study was associated with hypoferritinemia and a significant decrease in serum iron compared to NAP (*p* <0.001). Several authors also report these characteristics in anemia and iron deficiency [11,44,45]. In fact, iron deficiency develops in two major stages. The first stage corresponds to a reduction in the body’s iron reserves without abnormalities in the blood count [46]. At this stage, the most sensitive routine test is the measurement of ferritin levels, which in this case is lower than the usual values [47]. The second stage corresponds to a depletion of iron reserves which this time has an impact on erythropoiesis. This impact will initially be reflected on the blood count by a decrease in the mean corpuscular hemoglobin content (MCHC) (hypochromia), and in the mean corpuscular volume of the red blood cell (MCV) [48] (microcytosis), then secondarily, by anemia if siderode erythropoiesis continues. Indeed, the defect in hemoglobin synthesis induces an intramedullary abortion of erythroblasts. The reticulocyte count is aregenerative in nature. Classically, isolated iron deficiency anemia is called hypochromic aregenerative microcytic [48]. According to Cook, there are two major mechanisms leading to iron-deficient erythropoiesis: iron deficiency anemia and pathologies interfering with iron metabolism, namely chronic inflammatory or neoplastic diseases [48].

One of the strengths of this study is that it provides explanations on the characteristics of anemia and iron deficiency anemia in diabetic patients. One of the limitations lies in the fact that it was carried out in analytical cross-sectional; all the patients included were diabetic; a comparison of the results obtained with those of a non-diabetic population would have allowed to better understand the mechanism and characteristics of anemia and iron deficiency in diabetics. On the other hand, an exploration of the extended iron balance would allow to have complete information on the iron balance in patients.

## 5. CONCLUSION

The aim of this study was to biologically characterize anemia and iron deficiency anemia and identify the factors associated with the latter in diabetic patients at the Bafoussam Regional Hospital in West Region of Cameroon. This study showed a high frequency of anemia (64.3%); of which 11.4% were iron deficiency anemia. A large proportion of the patients studied had hyperglycemia and high HbA1c. The blood count revealed hematological abnormalities characterized by leukopenia, granulocytopenia, thrombocytopenia with macroplateletosis. The anemia was characterized by microcytosis, hypochromia and significant regeneration. Iron deficiency was associated with hypoferritinemia, low serum iron. Macroplatelets and high Hb1AC were identified as predictive factors of iron deficiency anemia in diabetes with respectively (OR=2.83, 95% CI=[1.17;6.80]; *p*=0.021) and OR=2.38, 95% CI=[0.14;3.09], *p*=0.045. This study raises the need for regular exploration of iron status in diabetics to prevent complications associated with iron deficiency.

## Data Availability

All relevant data are within the manuscript and its Supporting Information files.

## Funding

This research did not receive any specific grant from funding agencies in the public, commercial, or not-for-profit sectors.

## Conflicts of Interest

The authors declare that they have no conflicts of interest regarding the publication of this manuscript.

## Authors’ Contributions

Melisa Mene Djeukamgang carried out the data collection, and analysis of the biological samples and drafted the manuscript. Romaric Tuono De Manfouo, Maï-Pamela Tumchou Mekieje and Josué Louokdom Simo interpreted the data and contributed to the drafting of the manuscript. Pascal Blaise Well a Well a Koul, and Marius Mbiandjeu Tchoumke contributed to the analysis and interpretation of the data, and the redaction of the manuscript. Dieudonne Pascal Chuisseu Djamen, Josué Louokdom Simo and Leonard Tedong Josué supervised the work. All authors read and approved the final version of the manuscript, and Romaric Tuono De Manfouo had full access to all of the data in this study and took complete responsibility for the integrity of the data and the accuracy of the data analysis.

## Acknowledgments

We offer our sincere thanks to all the staff of laboratories of the “*Hôpital Regional de Bafoussam”*, and “*Université des Montagnes”* for their help in carrying out this work.

## Transparency Statement

The lead author Romaric Tuono De Manfouo affirms that this manuscript is an honest, accurate, and transparent account of the study being reported; that no important aspects of the study have been omitted; and that any discrepancies from the study as planned (and, if relevant, registered) have been explained".

## Data Availability

The authors confirm that the data supporting the findings of this study are available from the corresponding author.

## Notes

### Competing Interest Statement

The authors have declared no competing interest.

### Funding Statement

The author(s) received no specific funding for this work.

### Author Declarations

This study was conducted after obtaining study authorizations and ethical clearance issued by the Institutional Ethics and Research Committee of the University of the Mountains (No. 2024/211/UdM/PR/CEAQ). The authorization for collection and analysis (No. 663/L/MINSANTE/SG/DRSPO/HRB/D) was obtained by the Regional Hospital of Bafoussam.

